# Evaluation of Hi-C sequencing for the detection of gene fusions in hematologic and solid pediatric cancer samples

**DOI:** 10.1101/2024.05.10.24306838

**Authors:** Anthony D. Schmitt, Kristin Sikkink, Atif A. Ahmed, Shadi Melnyk, Derek Reid, Logan Van Meter, Erin M. Guest, Lisa A. Lansdon, Tomi Pastinen, Irina Pushel, Byunggil Yoo, Midhat S. Farooqi

## Abstract

HiC sequencing is a DNA-based next-generation sequencing method that preserves the 3D conformation of the genome and has shown promise in detecting genomic rearrangements in translational research studies. To evaluate HiC as a potential clinical diagnostic platform, analytical concordance with routine laboratory testing was assessed using primary pediatric leukemia and sarcoma specimens previously positive for clinically significant genomic rearrangements. Archived specimen types tested included viable and nonviable frozen leukemic cells, as well as formalin-fixed paraffin-embedded (FFPE) tumor tissues. Initially, pediatric acute myeloid leukemia (AML) and alveolar rhabdomyosarcoma (A-RMS) specimens with known genomic rearrangements were subjected to HiC analysis to assess analytical concordance. Subsequently, a discovery cohort consisting of AML and acute lymphoblastic leukemia (ALL) cases with no known genomic rearrangements based on prior clinical diagnostic testing were evaluated to determine whether HiC could detect rearrangements. Using a standard sequencing depth of 50 million raw read-pairs per sample, or approximately 5X raw genomic coverage, 100% concordance was observed between HiC and previous clinical cytogenetic and molecular testing. In the discovery cohort, a clinically relevant gene fusion was detected in 45% of leukemia cases (5/11). This study demonstrates the value of HiC sequencing to medical diagnostic testing as it identified several clinically significant rearrangements, including those that might have been missed by current clinical testing workflows.

**Key points:**

- HiC sequencing is a DNA-based next-generation sequencing method that preserves the 3D conformation of the genome, facilitating detection of genomic rearrangements.
- HiC was 100% concordant with clinical diagnostic testing workflows for detecting clinically significant genomic rearrangements in pediatric leukemia and rhabdomyosarcoma specimens.
- HiC detected clinically significant genomic rearrangements not previously detected by prior clinical cytogenetic and molecular testing.
- HiC performed well with archived non-viable and viable frozen leukemic cell samples, as well as archived formalin-fixed paraffin-embedded tumor tissue specimens.

## Introduction

Pediatric cancer is the leading cause of disease-induced morbidity and mortality in children in the USA and encompasses a wide range of cancer types, including leukemias, lymphomas, and sarcomas.^1^ Advances in sequencing technologies have contributed to a better understanding of the molecular characteristics of these diseases and have provided clues to new markers for targeted therapy.^2, 3^

While the mutational burden of pediatric cancers is generally lower than their respective adult counterparts^4, 5^, genomic rearrangements, including gene fusions and other chromosomal rearrangements, are more common drivers in pediatric cancer.^3, 5, 6^ These genomic rearrangements can result in the overexpression or loss-of-function of genes, or create entirely new chimeric genes, which may then serve as biomarkers for disease, give information as to prognosis, and/or be therapeutically targetable.

Clinical testing methods for genomic rearrangements are shared across pediatric cancer types, including rhabdomyosarcomas and leukemias; commonly used techniques include cytogenetic methods such as karyotyping, fluorescence *in situ* hybridization (FISH), and chromosomal microarray analysis (CMA). However, analysis of genomic rearrangements in pediatric cancer using these routine cytogenetic testing methods is complicated by several factors. First, live cells must be obtained and grown in culture for karyotyping. This is often not possible for solid tumors where tissue is formalin-fixed and paraffin-embedded (FFPE) and a fresh sample was not set aside for karyotyping analysis, or if karyotyping failed due to poor tumor cell growth in culture.^7^ Furthermore, karyotype analysis has a relatively coarse genetic resolution, detecting only structural changes greater than 5-10Mb, rendering some rearrangements cryptic and undetectable.^8^ Second, FISH requires the selection of probes for the assay and therefore cannot detect genomic rearrangements involving novel genes, or, depending on the FISH probe design (breakapart vs. dual fusion), cannot resolve the fusion partner or detect fusions involving novel (unprobed) partners, respectively. FISH also can be fraught with a higher background signal when analyzing FFPE solid tumor tissue potentiating false negative or false positive calls.^9^ Third, CMA is unable to detect balanced genomic rearrangements, which are important known drivers of pediatric cancers.^10^ There are also notable strengths of cytogenetic techniques, including relatively fast turnaround time and low limit of detection, as both karyotype and FISH are inherently single-cell resolution techniques. However, these strengths are counterbalanced by operational complexities, requiring specialized lab and interpretation expertise to conduct FISH and karyotype analyses limiting the number of clinical laboratories that can perform such analyses.^11^

Molecular methods for detecting gene fusions, such as targeted RNA sequencing (RNA-seq) approaches, including anchored multiplex PCR (AMP), are increasingly utilized for detecting gene fusions in solid and hematological cancers.^12–15^ However, for solid tumors stored as FFPE samples, RNA degradation can preclude reliable analysis in routine diagnostic use; this issue is further exacerbated for research specimens with longer archival periods.^16, 17^ Also, even though AMP is partner-agnostic, it still requires targeting of at least one gene involved in a given gene fusion, thereby missing fusions where both genes are untargeted.^15^ More generally, AMP is also insensitive to other types of genomic rearrangements, such as non-coding rearrangements (promoter swaps or enhancer hijacking events) that do not produce fusion transcripts, which are routinely tested for and inform clinical decision-making in hematological cancers, such as acute lymphoblastic leukemia (ALL).^18^ Other molecular methods include optical genome mapping (OGM). Since the benefits of OGM are dependent upon the successful isolation of high molecular weight (HMW) DNA, OGM has been deployed by some clinical labs for hematological cancers where HMW DNA can be isolated and preserved from higher quality specimens. However, OGM currently cannot be applied to FFPE tissues where the DNA is potentially already fragmented and/or the tissue specimen quality is compromised, thus precluding OGM analyses of FFPE solid tumors.^19^

In this study, we evaluated a 3D genomics approach to detect genomic rearrangements in primary pediatric leukemia and sarcoma specimens. The approach, termed HiC sequencing, is a DNA-based next-generation sequencing (NGS) method that preserves the 3D conformation of the genome. HiC sequencing has emerged as a promising approach for genomic rearrangement detection in translational research studies and has been previously shown to be an accurate and sensitive approach for identifying genomic rearrangements, including inter- and intra-chromosomal translocations and other structural rearrangements.^20–29^ However, detailed evaluations of the technology with a focus on analytical concordance relative to routine clinical laboratory testing workflows in solid and hematological cancers are lacking, as is an evaluation of how HiC-based testing could impact the diagnostic yield by detecting clinically significant gene fusions, either in the context of supplementing or replacing technologies in current clinical testing workflows. Overall, this study aimed to provide an institutional proof of principle evaluation of HiC for genomic rearrangement detection in pediatric cancer and sought to determine its utility in clinical testing workflows.

## Materials and Methods

### Overview

First, to evaluate analytical concordance, archived pediatric acute myeloid leukemia (AML) and alveolar rhabdomyosarcoma (A-RMS) specimens with known gene fusions as determined by prior clinical testing were subjected to Arima Genomics’ HiC sample prep, followed by Illumina sequencing and genomic rearrangement bioinformatics analysis. Next, to explore the potential impact on diagnostic yield using HiC, a discovery cohort with additional AML and ALL specimens were analyzed. These specimens had been previously subjected to standard-of-care clinical cytogenetic and/or molecular testing, and no genetic driver/known gene fusion had been identified. Hematologic specimens included frozen white blood cell pellets and viably preserved frozen hematopoietic cells, either from bone marrow aspirates or peripheral blood. For solid tumor specimens, unstained FFPE scrolls were used.

### Samples – Solid Tumors

Pediatric A-RMS specimens were collected by the Children’s Mercy Hospital (CMH, Kansas City, MO, USA) as part of the routine diagnostic workflow. Each specimen was formalin-fixed and paraffin embedded and stored at room temperature. FFPE A-RMS samples (N=5, archival period range 9-13 years) that were known to be gene fusion-positive as detected by previous clinical testing (i.e., chromosomes, FISH, or microarray) were sectioned to produce unstained tissue scrolls (n=3 to 5 scrolls per sample) for HiC library preparation and sequencing. The HiC workflow and genetic analyses were performed blinded to the clinical genetic results. A summary of the sample types, preservation methods, and archival periods is provided in **Supplemental Table S1**. A retrospective study, including genetic analysis, of deidentified archived paraffin-embedded RMS tissue was approved by the CMH Institutional Review Board (IRB).

### Samples – Leukemias

Leukemia specimens were collected by CMH into its Biorepository. This was done under a Tumor Bank research study that included patient consent, and collection, processing, and storage of patient samples, as approved by the CMH IRB. Specimens collected prior to 2019 underwent white blood cell (WBC) isolation, were snap-frozen (non-viably), and stored at – 80°C. Specimens collected after 2019 underwent WBC isolation and were gradually frozen (viably) in a preservation media comprising 10% DMSO and tissue culture media, and then stored at –80°C.

For the concordance cohort, five AML specimens were cryopreserved, while two were frozen non-viably (N=7). Six AML samples were obtained from bone marrow aspirates, while one was a peripheral blood specimen (N=7 total; archival period range 1-4 years, **Supplemental Table S1**). In the concordance cohort, each specimen had undergone clinical standard-of-care cytogenetic (karyotyping, FISH, and/or microarray) and/or molecular testing (targeted cancer NGS sequencing panel, PCR). For five of the specimens, a known gene fusion had been identified. For two specimens, no known gene fusions had been identified and instead driver alterations other than gene fusions had been detected (e.g., biallelic *CEBPa* mutations). Once again, the HiC workflow and genetic analyses were performed blinded to the clinical genetic results.

For the discovery cohort, eight leukemia specimens were cryopreserved, while three specimens were frozen non-viably (N=11). Ten specimens were bone marrow aspirates, and one was a peripheral blood specimen (N=11 total), comprising precursor B-ALL, T-ALL, and AML. Archival period ranged from 1-4 years. In the discovery cohort, each specimen had undergone clinical standard-of-care cytogenetic (karyotyping, FISH, microarray) and/or molecular testing (targeted cancer NGS sequencing panel) and no genetic driver or known gene fusion had been identified. For all leukemia specimens, a summary of the sample types, preservation methods, and archival periods is provided in **Supplemental Table S1**.

### Arima-HiC sequencing

For frozen cell pellets or viably-preserved cells isolated from blood, cells were first crosslinked and then subjected to HiC sample preparation using the Arima-HiC kit (P/N A51008, Arima Genomics, Carlsbad, CA). For FFPE tissues, 5-10 μm sections were first de-waxed and rehydrated and then subjected to HiC sample preparation using the Arima-HiC for FFPE kit (P/N A311038). Subsequently, short-read sequencing libraries were prepared by shearing the proximally ligated DNA, followed by size-selecting DNA fragments using solid phase reversible immobilization (SPRI) beads. DNA fragments containing ligation junctions were then enriched using Enrichment Beads (provided in the Arima-HiC kits) and converted into sequencing libraries using the Swift Accel-NGS 2S Plus kit (Swift Biosciences, P/N: 21024). After adapter ligation, DNA was PCR amplified and purified using SPRI beads. The purified DNA underwent standard quality control (qPCR and Bioanalyzer) and was sequenced using a NovaSeq 6000 (Illumina, San Diego, CA) according to the manufacturer’s instructions.

### Data analysis workflow

Genomic rearrangements were identified using the Arima-SV v1.3 pipeline.^30^ The Arima-SV v1.3 pipeline contains several sub-components. First, raw read-pairs are aligned to the human reference genome (hg38) and deduplicated using HiCUP^31^. Genomic rearrangements are called using HiC-Breakfinder.^20^ For data visualization, de-duplicated alignments from HiCUP are converted into HiC matrices using Juicer software^32^, which are then visualized alongside the genomic rearrangement calls using Juicebox.^33^ For sub-sampling analyses, raw read-pairs were randomly extracted from the full datasets using a sub-sampling feature of the Arima-SV v1.3 pipeline, and then the sub-sampled raw read-pairs were run through the Arima-SV v1.3 workflow for rearrangement detection and data visualization.

## Results

### HiC library preparation and quality assessment

Arima-HiC libraries prepared from retrospective leukemia and sarcoma cases were deeply sequenced, and genomic rearrangements were detected using the Arima-SV v1.3 pipeline (**Figure 1**). Each full sequencing dataset was evaluated for quality using well established performance metrics for HiC data that are output from the Arima-SV v1.3 pipeline (**Supplemental Table S2**). Of note, within the leukemia specimens, we observed that the data quality for the specimens prepared as frozen non-viable cell pellets was considerably lower than the specimens prepared as frozen viable (cryopreserved) cells. This is most likely due to cells bursting upon non-viable freezing but may also be exacerbated by the archival periods (e.g., 4 years) or other factors. Also of note, the FFPE specimens with an archival period of 9-13 years had relatively low DNA yields and reduced data quality metrics relative to FFPE specimens with shorter archival periods. Lastly, while data quality was assessed using the original full sequencing depths (**Supplemental Table S2)**, we performed all genomic rearrangement analyses in the leukemia and sarcoma specimens at a sub-sampled raw sequencing depth of 50 million raw read-pairs (∼5X raw genome coverage). The sub-sampled datasets had quality metrics resembling that of the full depth datasets except with fewer mapped HiC read-pairs available for analysis.

**Figure 1.**
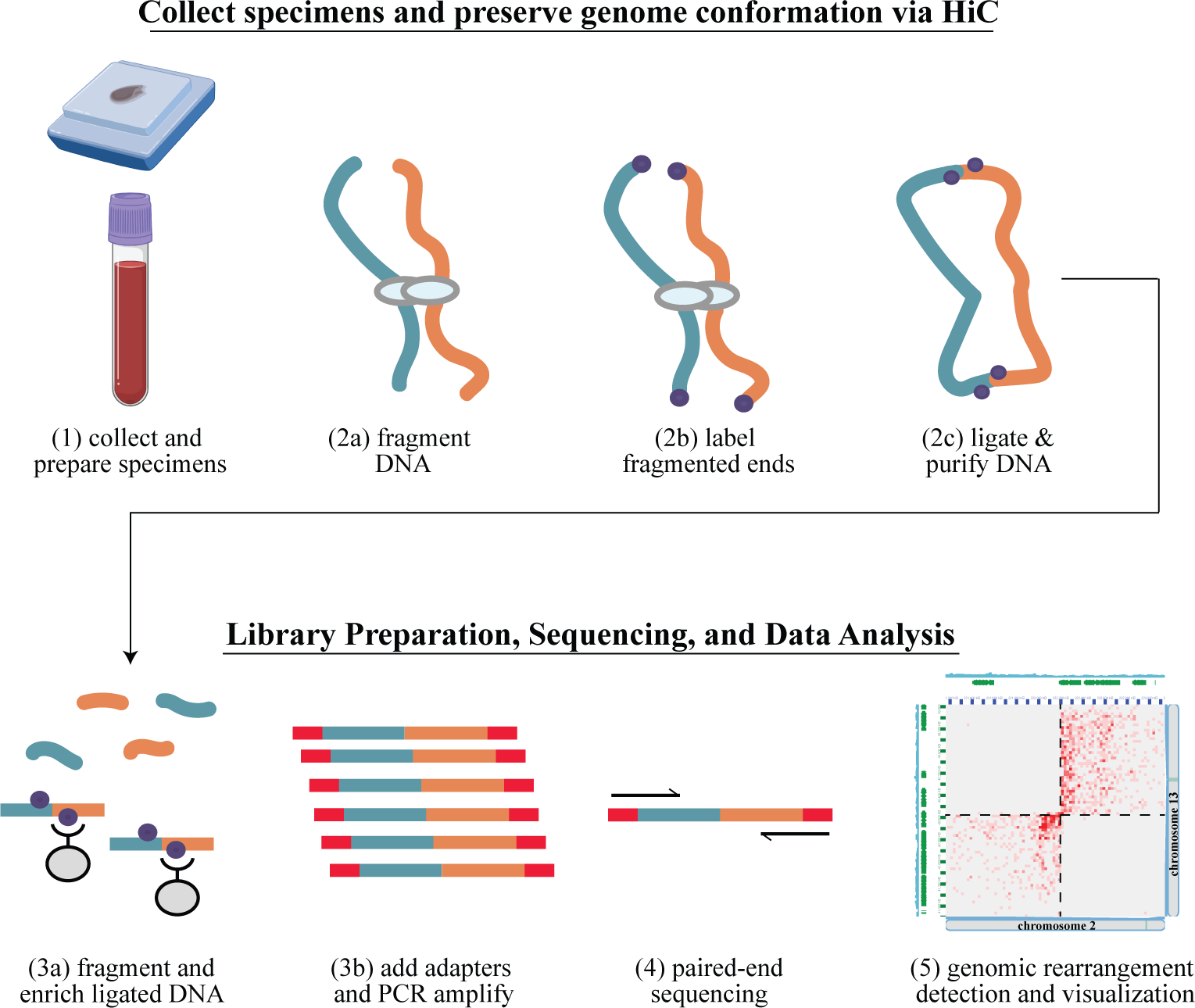
Genomic rearrangement detection using Arima Genomics’ HiC workflow. (Step 1) collect specimens, and prepare for HiC testing. For hematologic cancers, extract the white blood cells from the bone marrow aspirates or peripheral blood and crosslink. For formalin-fixed paraffin-embedded (FFPE) solid tumors, de-wax and rehydrate the tissue; (Step 2) preserve the 3D conformation of the genome via HiC, resulting in labeled proximity ligated DNA that has preserved 3D conformation information; (Step 3) library preparation, resulting in a sequence-ready HiC library; (Step 4) next-generation sequencing (NGS); (Step 5) bioinformatics analysis using the Arima-SV v1.3 workflow to identify genomic rearrangements and visualize results.

### HiC is concordant with gold-standard methods for pediatric cancer clinical cytogenetic testing

We used the sub-sampled datasets to 50 million raw read-pairs to determine concordance with clinical cytogenetic testing. At this depth, the clinically relevant gene fusions in all five previously determined fusion-positive AML cases were detected (two *RUNX1::RUNX1T1* fusions, one *CBFB::MYH11* fusion, one *CBFA2T3::GLIS2* fusion, and one *KMT2A*::*MLLT4* fusion) (**Table 1)**. For example, in the “AML C6” case, prior clinical testing from karyotype analysis showed an apparent chromosome 11 deletion and translocation with the long arm of chromosome 6. A panel of FISH assays found a *KMT2A* rearrangement with *3’KMT2A* loss, and microarray analysis found deletions of *KMT2A* exons 11-36 and *MLLT4* exon 1 deletion, suggestive of a *KMT2A*::*MLLT4* fusion. HiC analysis detected a genomic rearrangement between chromosome 11 and the long arm of chromosome 6, but also the reciprocal rearrangement when a telomeric portion of chromosome 6 translocated with the long-arm of chromosome 11. These findings are apparent when the HiC data is viewed at the chromosome-scale (**Figure 2A**), where the “bowtie” signal pattern in the HiC data is known to be indicative of a reciprocal inter-chromosomal rearrangement.^27^ Upon inspection of the genomic rearrangement breakpoint at higher resolution, the *KMT2A*::*MLLT4* gene fusion is readily detected, as well as apparent reductions in sequencing coverage at both the 3’ portion of *KMT2A* and the 5’ portion of *MLLT4* (**Figure 2B**). The HiC results were further supported by conventional NGS-based testing done at a commercial reference laboratory, which also detected the *KMT2A::MLLT4* gene fusion.

**Figure 2.**
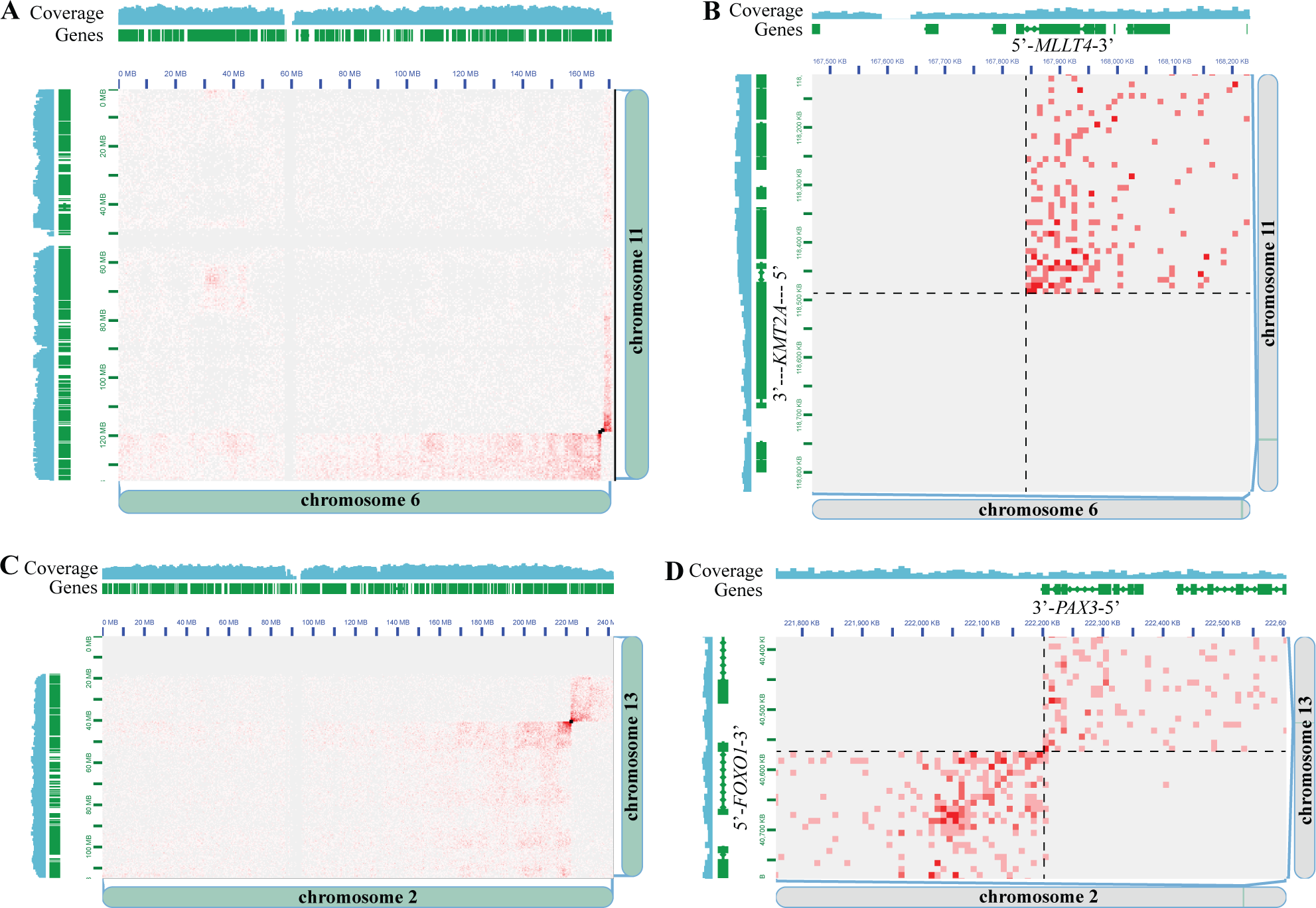
HiC is concordant with clinical cytogenetic testing for detecting clinically significant gene fusions. A) Chromosome 6 x Chromosome 11 HiC heatmap from specimen “AML C6”. Genomic coordinates, gene locations, and sequencing coverage from Chromosome 6 and 11 are shown along the edges of the X and Y axes of the heatmap, respectively. Small black boxes overlaid on the heatmap are the Arima-SV pipeline genomic rearrangement calls. B) Same as panel A, except zoomed-in to the locus around the *KMT2A::MLLT4* gene fusion call. The *KMT2A* and *MLLT4* gene positions and orientations are indicated. Black dashed lines depict the breakpoint locations on each chromosome. C) Chromosome 2 x Chromosome 13 HiC heatmap from specimen “ARMS C3”. Genomic coordinates, gene locations, and sequencing coverage from Chromosome 2 and 13 are shown along the edges of the X and Y axes of the heatmap, respectively. Small black boxes overlaid on the heatmap are the Arima-SV pipeline genomic rearrangement calls. D) Same as panel C, except zoomed-in to the locus around the *PAX3::FOXO1* gene fusion call. The *PAX3* and *FOXO1* gene positions and orientations are indicated. Black dashed lines depict the breakpoint locations on each chromosome.

**Table 1:** HiC is concordant with clinical cytogenetic testing.

| Sample ID | Cancer Type | Sample Source | Preservation (if any) | Prior Cytogenetic Test Result | HiC Test Result |
| --- | --- | --- | --- | --- | --- |
| AML C1 | AML | PBMCs | 90% FBS/10% DMSO | *CBFA2T3::GLIS2* | *CBFA2T3::GLIS2* |
| AML C2 | AML | Bone Marrow | None | Negative* | Negative |
| AML C3 | AML | Bone Marrow | None | *RUNX1::RUNX1T1* | *RUNX1::RUNX1T1* |
| AML C4 | AML | Bone Marrow | 90% FBS/10% DMSO | *MYH11::CBFB* | *MYH11::CBFB* |
| AML C5 | AML | Bone Marrow | 90% FBS/10% DMSO | Negative** | Negative |
| AML C6 | AML | Bone Marrow | 90% FBS/10% DMSO | *KMT2A::MLLT4\*\*\** | *KMT2A::MLLT4* |
| AML C7 | AML | Bone Marrow | 90% FBS/10% DMSO | *RUNX1::RUNX1T1* | *RUNX1::RUNX1T1* |
| ARMS C1 | A-RMS | Tissue | FFPE | *FOXO1-r* | *PAX3::FOXO1* |
| ARMS C2 | A-RMS | Tissue | FFPE | *PAX3::FOXO1* | *PAX3::FOXO1* |
| ARMS C3 | A-RMS | Tissue | FFPE | *PAX3::FOXO1* | *PAX3::FOXO1* |
| ARMS C4 | A-RMS | Tissue | FFPE | *FOXO1-r* | *PAX7::FOXO1* |
| ARMS C5 | A-RMS | Tissue | FFPE | *FOX7::FOXO1* | *PAX7::FOXO1* |

*AML = Acute Myeloid Leukemia; A-RMS = Alveolar Rhabdomyosarcoma; PBMC = Peripheral Blood Mononuclear Cell; FBS = Fetal Bovine Serum; DMSO = Dimethyl Sulfoxide; FFPE = Formalin-Fixed Paraffin-Embedded; “-r” indicates when the fusion partner is unknown.*
*\* FLT3 ITD mutation detected (PCR)*
*\*\* Biallelic CEBPa mutations detected (targeted DNA-seq)*
*\*\*\* KMT2A::MLLT4 fusion also detected upon NGS-based fusion testing*

In addition, the clinically relevant gene fusions in all five previously determined fusion-positive A-RMS cases were detected (*PAX3::FOXO1* (N=3) and *PAX7::FOXO1* (N=2)) (**Table 1**). For example, in the “ARMS C3” case, previous clinical testing from FISH analysis showed evidence for *FOXO1* (FHKR) gene rearrangement and aneuploidy, and karyotype analysis showed t(2;13) indicative of *PAX3*::*FOXO1* gene fusion. HiC analysis detected a reciprocal genomic rearrangement between chromosomes 2 and 13 (**Figure 2C**), and upon inspection at the genomic rearrangement breakpoint at higher resolution, the *PAX3*::*FOXO1* gene fusion was readily detected (**Figure 2D**). Furthermore, because the signal pattern in the HiC data is indicative of the type of chromosomal rearrangement, HiC analysis can distinguish between chromosomal rearrangements and extrachromosomal rearrangements such as double minutes.^34–38^ Analysis of the HiC data predicted the chromosomal rearrangement type as double minutes for three of five A-RMS cases, and as chromosomal translocations for two of five A-RMS cases. These predictions agreed with the karyotype analysis for all four cases where karyotype analysis was performed (**Supplemental Figure S1**).

While these concordance analyses were all carried out at a standardized sequencing depth of 50 million raw read-pairs, we also determined the minimum sequencing depths at which the clinically relevant gene fusions could be detected. In the FFPE sarcoma samples, 5/5 (100%) gene fusions could be detected at 25 million raw read-pairs, 4/5 (80%) at 12.5 million raw read-pairs, and 3/5 (60%) at 5 million raw read-pairs (**Supplemental Table S3**). Furthermore, in the AML specimens, 4/5 (80%) gene fusions could be detected at 25 and 12.5 million raw read-pairs, and 3/5 (60%) at 5 million raw read-pairs (**Supplemental Table S3**). The one AML specimen insensitive at 25 million and 12.5 million raw reads was the only specimen amongst the fusion-positive AML concordance cases prepared as a non-viable frozen pellet, which had lower HiC data quality metrics, indicating an interplay between HiC data quality profiles and analytical sensitivity that requires further systematic analysis. Unsurprisingly, no fusions were detected across all sequencing depths in the two fusion-negative AML specimens in the concordance cohort.

### Driver-negative leukemias – Discovery cohort

We then assessed a discovery cohort of leukemia specimens for which previous diagnostic testing had not identified any driver genomic rearrangements. Samples included one AML, eight B-ALL, and two T-ALL specimens (**Table 2, Supplemental Table 2**). Similar to the concordance cases, we analyzed the data at a standardized depth of 50 million raw read-pairs. We detected clinically significant fusions in 3 of 11 cases (two *ZNF384::EP300* fusions and one *KMT2A::MLLT10* fusion), as well as potentially significant fusions in 2 of 11 cases (one *SKAP2::CDK6* fusion, and one *ABHD17B::PTK2B* fusion) (**Table 2**). For example, in the “AML D1” case, HiC analysis detected a *KMT2A*::*MLLT10* gene fusion (**Figure 3A,B)**, while previous clinical testing comprising of karyotyping, FISH, short-read NGS, and microarray did not detect this clinically significant rearrangement. This observation is consistent with reports of this rearrangement sometimes being cryptic and not visible on karyotype analysis, FISH, or microarray.^39^ Indeed, interpretation of the HiC signal pattern at the *KMT2A::MLLT10* breakpoint (**Figure 3B**) indicates an inversion of the 5’ portion of *KMT2A,* and fusion to the 3’ portion of *MLLT10*, resulting in the expected *KMT2A::MLLT10* fusion gene orientation. Identification of the *KMT2A*::*MLLT10* gene fusion carries strong clinical significance; had it been detected in the prospective setting, it would have changed the pathology diagnosis, risk classification, and treatment approach.^40^ *KMT2A*::*MLLT10* is considered a high-risk fusion in pediatric AML and is an indication for hematopoietic stem cell transplant in first remission.^41, 42^ There is also evidence that addition of Gemtuzumab to conventional chemotherapy improves clinical outcomes for patients with *KMT2A*-rearranged AML.^41^ In two other B-ALL cases (“B-ALL D8” and “B-ALL D6”), HiC analysis detected *ZNF384::EP300* gene fusions (**Figure 3C-F)**, which previous clinical testing comprising of karyotyping, FISH, short-read NGS, and microarray did not detect (**Table 2**). This finding also carries clinical significance, whereby had the *ZNF384*::*EP300* gene fusions been detected in the prospective setting, it would have changed the pathology diagnosis^40^ and been associated with an intermediate prognosis. In addition, patients with *ZNF384* fusions to *EP300* have been reported to have lower relapse rates than patients with other *ZNF384* fusions.^43^ Of note, this rearrangement is known to be cytogenetically cryptic, was not specifically tested for by FISH, and is balanced so was not detected by CMA. Furthermore, RNA-based fusion panel testing was performed at a reference laboratory for case B-ALL D8 but was negative, consistent with the panel not targeting *EP300* or *ZNF384*. This scenario underscores the utility of HiC given its unbiased genome-wide nature for fusion detection with relatively low-pass coverage (∼50 million raw read-pairs).

**Figure 3.**
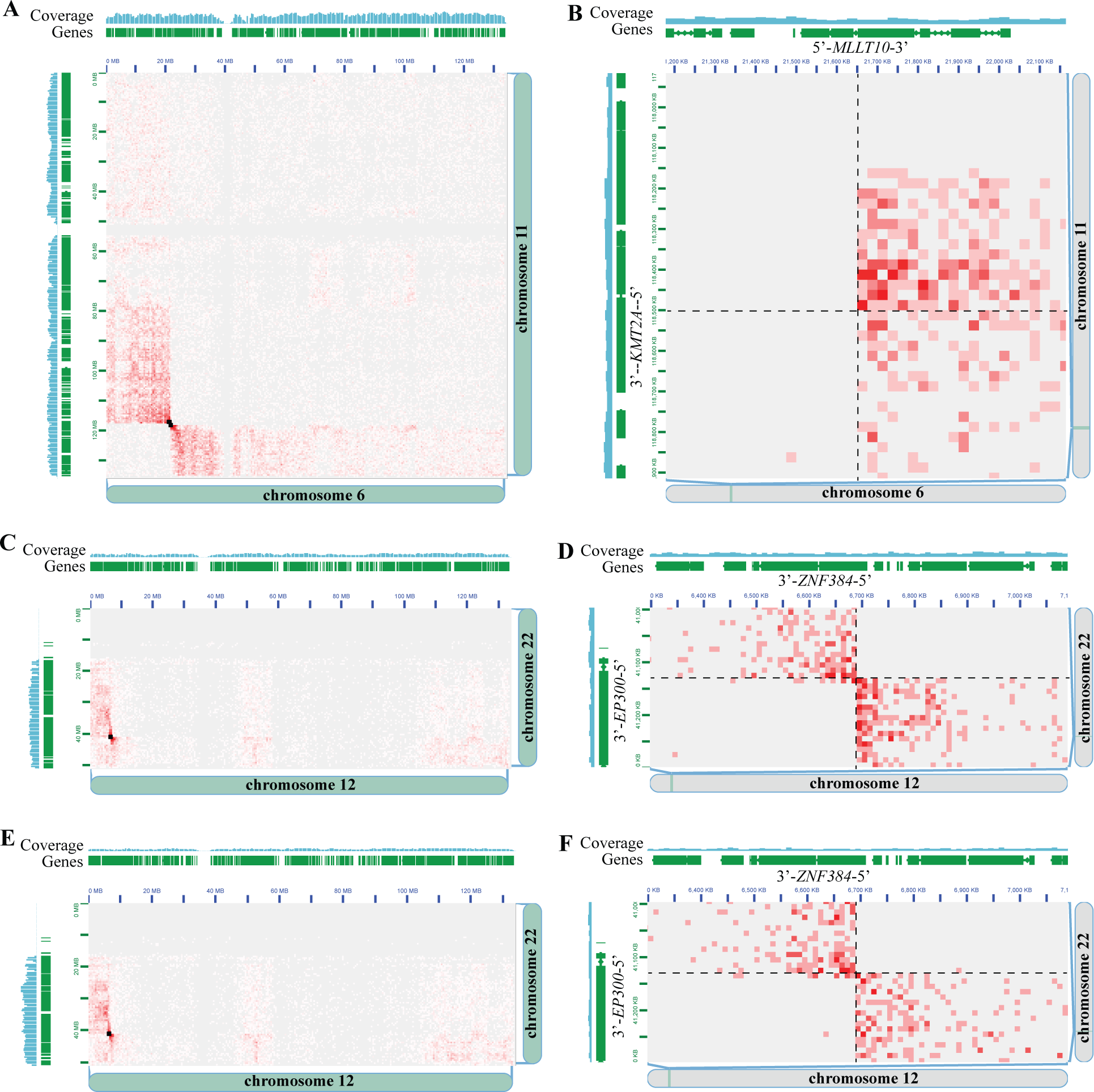
HiC detects clinically significant gene fusions not previously detected by clinical cytogenetic and molecular testing. A) Chromosome 6 x Chromosome 11 HiC heatmap from specimen “AML D1”. Genomic coordinates, gene locations, and sequencing coverage from Chromosome 6 and 11 are shown along the edges of the X and Y axes of the heatmap, respectively. Small black boxes overlaid on the heatmap are the Arima-SV pipeline genomic rearrangement calls. B) Same as panel A, except zoomed-in to the locus around the *KMT2A::MLLT10* gene fusion call. The *KMT2A* and *MLLT10* gene positions and orientations are indicated. Black dashed lines depict the breakpoint locations on each chromosome. C) Chromosome 12 x Chromosome 22 HiC heatmap from specimen “B-ALL D8”. Genomic coordinates, gene locations, and sequencing coverage from Chromosome 12 and 22 are shown along the edges of the X and Y axes of the heatmap, respectively. Small black boxes overlaid on the heatmap are the Arima-SV pipeline genomic rearrangement calls. D) Same as panel C, except zoomed-in to the locus around the *ZNF384::EP300* gene fusion call. The *ZNF384* and *EP300* gene positions and orientations are indicated. Black dashed lines depict the breakpoint locations on each chromosome. E) Same as panel C, except from specimen “B-ALL D6”. F) Same as panel D, except from specimen “B-ALL D6”.

**Table 2.** HiC detects clinically significant gene fusions not detected by prior clinical cytogenetic and molecular testing.

| Sample ID | Cancer Type | Sample Source | Preservation (if any) | Prior Dx Test Result | HiC Test Result | Clinical Impact |
| --- | --- | --- | --- | --- | --- | --- |
| AML D1 | AML | Bone Marrow | 90% FBS/10% DMSO | Negative | *KMT2A::MLLT10* | Dx, Px, Tx |
| T-ALL D1 | T-ALL | Bone Marrow | 90% FBS/10% DMSO | Negative | *SKAP2::CDK6* | Tx* |
| T-ALL D2 | T-ALL | Bone Marrow | None | Negative | Negative | N/A |
| B-ALL D1 | B-ALL | Bone Marrow | None | Negative | Negative | N/A |
| B-ALL D2 | B-ALL | Bone Marrow | None | Negative | Negative | N/A |
| B-ALL D3 | B-ALL | Bone Marrow | 90% FBS/10% DMSO | Negative | Negative | N/A |
| B-ALL D4 | B-ALL | Bone Marrow | 90% FBS/10% DMSO | Negative | Negative | N/A |
| B-ALL D5 | B-ALL | PBMCs | 90% FBS/10% DMSO | Negative | Negative | N/A |
| B-ALL D6 | B-ALL | Bone Marrow | 90% FBS/10% DMSO | Negative | *ZNF384::EP300* | Dx, Px |
| B-ALL D7 | B-ALL | Bone Marrow | 90% FBS/10% DMSO | Negative | *ABHD17B::PTK2B* | Dx, Px** |
| B-ALL D8 | B-ALL | Bone Marrow | 90% FBS/10% DMSO | Negative | *ZNF384-EP300* | Dx, Px |

AML = Acute Myeloid Leukemia; ALL = Acute Lymphoblastic Leukemia; PBMC = Peripheral Blood Mononuclear Cell; FBS = Fetal Bovine Serum; DMSO = Dimethyl Sulfoxide; Dx = Diagnostic Clinical Impact; Px = Prognostic Clinical Impact; Tx = Therapeutic Clinical Impact; Clinical Impact is determined according to WHO and NCCN guidelines.
\* Phase I clinical trial for CDK4/6 inhibitors have been completed in pediatric and young adults with ALL<sup>45</sup>
\*\* PTK2Br is diagnostic and prognostic according to NCCN guidelines for ALL<sup>46</sup>

Similar to the analyses in the concordance cohort, we also determined the minimum sequencing depths at which the clinically relevant gene fusions could be detected. Across the five leukemia specimens with a clinically relevant gene fusion detected, 5/5 (100%) gene fusions could be detected at 25 and 12.5 million raw read-pairs, and 3/5 (60%) at 5 million raw read-pairs (**Supplemental Table S3**).

## Discussion

We conducted an institutional proof of concept evaluation of Arima Genomics’ HiC technology for the unbiased genome-wide detection of clinically relevant genomic rearrangements in a retrospective cohort of pediatric solid tumors and hematologic cancers. Using a standardized sequencing coverage of 50 million raw HiC read-pairs per sample (approximately 5X raw genomic coverage), we observed 100% (12/12) concordance between HiC and prior clinical genetic testing for clinically relevant gene fusions. Additionally, in pediatric leukemias without any previously detected genomic rearrangements via available clinical cytogenetic and molecular testing, HiC detected a clinically relevant gene fusion in ∼45% (5/11) of cases—providing diagnostically, prognostically, and therapeutically significant information.

One notable result from this study is that HiC is robust to a variety of clinical specimen types and preservation methods that are routinely obtained and processed in hematologic and solid tumor clinical and research laboratory testing workflows, and to various specimen preservation methods and archival periods. Importantly, for solid tumor clinical testing, HiC performed well using FFPE tissue, which represents a unique and powerful benefit to HiC technology compared to emerging long-read or optical mapping technologies whose advantages are dependent on the analysis of high-molecular-weight DNA, which is difficult to obtain from FFPE specimens. Furthermore, the FFPE specimens analyzed in this study had archival periods of 9 to 13 years. This suggests that in addition to prospective solid tumor testing, HiC technology opens up a range of research opportunities for pathologists or other clinical research investigators to study genomic rearrangements in archived FFPE tissue material. This is particularly important given that RNA-seq may perform poorly when analyzing FFPE specimens following prolonged storage due to RNA degradation.^16, 17^ In the hematological cancer setting, HiC detected clinically relevant genomic rearrangements in both viably and non-viably preserved specimens, although the data were of significantly better quality in viably preserved specimens. More systematic studies may be needed to tease apart the relationship between preservation method, HiC data quality, and fusion detection performance. Along those lines, this study also raises the need to define the compatibility of HiC in other sample types or preservation contexts that arise in clinical testing workflows for hematologic cancers. For example, compatibility with blood collected in blood collection tubes other than the ones used in this study (EDTA), blood processed after various time intervals between collection and HiC sample preparation (blood stability), and compatibility with other fixatives used in clinical testing workflows (e.g., acetic acid and methanol).

The genome-wide, partner-agnostic nature of the HiC approach utilized in this study has notable benefits for clinical testing compared to targeted approaches. For example, FISH requires the selection of probes for the assay and, depending on the FISH probe design, is potentially unable to resolve the fusion partner. FISH is also often performed serially as a single-gene test.^44^ In contrast, the genome-wide implementation of HiC can detect fusions involving all genes across the whole genome simultaneously and detects both partners for all fusions identified. However, FISH is inherently a technology with single-cell resolution and, therefore, capable of detecting fusions even when the percentage of tumor cells is very low, whereas the limit of detection for HiC remains to be defined.

Targeted RNA-seq has similar limitations since it is restricted to the list of genes on a given panel but can be multiplexed to hundreds of genes and can be partner-agnostic (e.g., AMP), assuming RNA quality and/or workflow complexity do not preclude RNA-seq analysis. While FISH and targeted RNA-seq can detect fusions involving genes already known (or suspected to be) of clinical relevance based on the probe/primer design, the genome-wide nature of HiC allows it to detect gene fusions of both known and unknown relevance, potentiating the discovery of novel biomarkers, disease mechanisms, or therapeutic targets to advance clinical research.

While this institutional proof of concept evaluation of Arima Genomics’ HiC technology for genomic rearrangement testing was successful, additional clinical validation studies would be needed to implement HiC into clinical laboratory workflows for either solid or hematological cancer testing. While these efforts lie ahead for academic clinical laboratories, Arima Genomics’ HiC technology is already available in the commercial clinical laboratory setting for genomic rearrangement analysis services (Aventa Genomics).

### Conclusion

Arima Genomics’ HiC technology was concordant with standard clinical diagnostic testing methods in solid and hematologic pediatric cancer specimens. Furthermore, the study demonstrated how HiC sequencing could provide additional diagnostic, prognostic, and therapeutic value by identifying clinically significant genomic rearrangements potentially missed by current clinical diagnostic testing workflows.

## Data Availability

Select non-identifying data in the present study are available upon reasonable request to the authors.

## Acknowledgements

Medical writing and editing services were provided by Maartje Wouters, which was funded by Arima Genomics. The authors would like to thank the patients and families who donated their samples for this research. The authors would also like to thank Lucia Laude and the Histology Core; Tumor Bank study personnel: Judy Vun, Amie Hatfield, and Robin Ryan; as well as Jason Seymour and Keiondra Sanders in the Children’s Mercy Research Institute Biorepository for their assistance with sample collection and processing. Finally, M.S.F. would like to thank Braden’s Hope for Childhood Cancer, Big Slick, and the Black & Veatch Foundation for their generous support of this work.

---

**Supplemental Figure S1. HiC analysis determines the rearrangement type from formalin-fixed paraffin-embedded solid tumors and is concordant with karyotype analysis.** The HiC signal pattern around the breakpoint calls inform the type of rearrangement. For simple inter-chromosomal translocations (translocations without co-occurring events at the breakpoint, such as inversions or deletions), there is a single breakpoint and the pattern of HiC signal strength will be strongest between pairs of loci directly flanking the breakpoints on each chromosome, and will dissipate for pairs of loci moving further away from the breakpoint. For simple reciprocal translocations, this looks like a bowtie pattern when viewing the loci around the breakpoint in a HiC heatmap, with strongest HiC signal strength at the center of the bowtie and with HiC signal strength dissipating bilaterally. For double minutes formed between genomic regions derived from two different chromosomes, the HiC signal pattern appears as a (often small) rectangle, with strongest HiC signal at opposing corners of the rectangle, and the HiC signal largely contained between the pair of genomic regions on the double minute. The Figure shows HiC heatmaps from the five alveolar rhabdomyosarcoma (ARMS) cases, zoomed-in to the loci surrounding the clinically significant gene fusion call. Genomic coordinates, gene locations, and sequencing coverage from each chromosome are shown along the edges of the X and Y axes of the heatmaps. The gene positions and orientations of the genes involved in the gene fusion are indicated. Black dashed lines depict the breakpoint location(s) on each chromosome. Cases “ARMS C1”, “ARMS C4”, and “ARMS C5” show the expected pair of breakpoints and HiC signal pattern of double minutes, whereas cases “ARMS C2” and “ARMS C3” show the expected HiC signal pattern of inter-chromosomal translocations.

