## Supplementary figures and images for "Evaluation of Hi-C sequencing for the detection of gene fusions in hematologic and solid pediatric cancer samples"

### Supplemental Figure S1

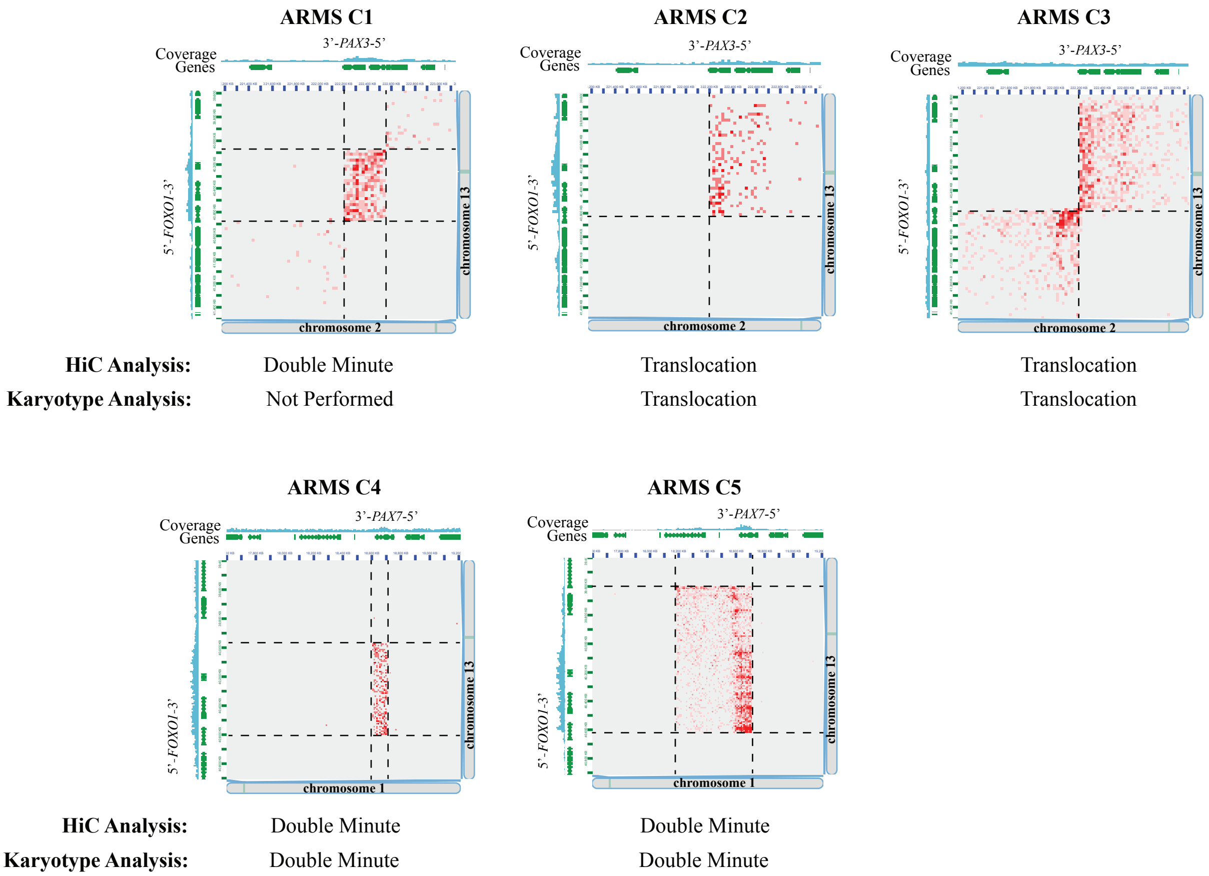
